# Humoral and Cellular Immunogenicity and Safety of 3 Doses of CoronaVac and BNT162b2 in Young Children and Adolescents with Kidney Diseases

**DOI:** 10.1101/2022.09.14.22279916

**Authors:** Daniel Leung, Eugene Yu-hin Chan, Xiaofeng Mu, Jaime S Rosa Duque, Samuel MS Cheng, Fanny Tsz-wai Ho, Pak-chiu Tong, Wai-ming Lai, Matthew HL Lee, Stella Chim, Issan YS Tam, Leo CH Tsang, Kelvin KH Kwan, Yuet Chung, Howard HW Wong, Amos MT Lee, Wing Yan Li, Summer TK Sze, Jennifer HY Lam, Derek HL Lee, Sau Man Chan, Wenwei Tu, Malik Peiris, Alison Lap-tak Ma, Yu Lung Lau

## Abstract

**Background:** Patients with kidney diseases are at risk of severe complications from COVID-19, yet little is known about the effectiveness of COVID-19 vaccines in children and adolescents with kidney diseases.

**Methods:** We investigated the immunogenicity and safety of an accelerated, 3-dose primary series of COVID-19 vaccines among 64 pediatric chronic kidney disease patients (mean age 12.2; 32 male) with or without immunosuppression, dialysis, or kidney transplant. CoronaVac was given to those aged <5 years, 0.1ml BNT162b2 to those aged 5-11 years, and 0.3ml BNT162b2 to those aged 11-18 years.

**Results:** Antibody responses including S-RBD IgG (90.9-100% seropositive) and surrogate virus neutralization (geometric mean sVNT% level, 78.6-94.0%) were significantly elicited by 3 doses of any vaccine. T cell responses were also elicited. Weaker neutralization responses were observed among kidney transplant recipients and non-dialysis children receiving rituximab for glomerular diseases. Neutralization was reduced against Omicron BA.1 compared to wild-type (post-dose 3 sVNT% level; 84% vs 27.2%; p<0.0001). However, T cell response against Omicron BA.1 was preserved, which likely confer protection against severe COVID-19. Hybrid immunity was observed after vaccination in infected patients, as evidenced by higher Omicron BA.1 neutralization response among infected patients receiving 2 doses than those uninfected. Generally mild or moderate adverse reactions following vaccines were reported.

**Conclusions:** Our findings support that an accelerated 3-dose primary series with CoronaVac and BNT162b2 is safe and immunogenic in young children and adolescents with kidney diseases.

**TRIAL REGISTRATION:** Clinicaltrials.gov NCT04800133

**SIGNIFICANCE STATEMENT:** Little is known about the effectiveness of COVID-19 vaccines in children and adolescents with kidney diseases. This paper describes the antibody and T cell responses of 3 doses of CoronaVac or BNT162b2, the top 2 COVID-19 vaccines distributed worldwide, by an accelerated regimen in patients with kidney diseases aged 1-18 years. Antibody and T cell responses were significantly elicited by either vaccine. Neutralization was reduced against Omicron while T cell response was preserved, which likely confer protection against severe COVID-19. Rate of severe adverse reactions was low in the study. Results confirm that accelerated 3-dose primary series with CoronaVac and BNT162b2 is safe and immunogenic in young children and adolescents with kidney diseases.

## INTRODUCTION

Patients with chronic kidney disease and kidney failure are at risk of severe disease and mortality in the COVID-19 pandemic.^1^ Impaired immune function in children and adolescents with severe kidney diseases predisposes them to infections,^2^ and vaccination offers protection to this vulnerable population. BNT162b2 is an mRNA vaccine with 90-100% efficacy against symptomatic COVID-19 caused by pre-Omicron variants in healthy children and adolescents aged 5-15 years.^3,4^ CoronaVac is a whole-virus inactivated vaccine with robust immunogenicity and real-world effectiveness demonstrated in children aged 3-17 years.^5-8^ In many places, including Hong Kong, immunocompromised children and adolescents are recommended to receive a 3-dose mRNA or inactivated vaccine primary series, due to their clinical vulnerability and suboptimal immunogenicity.^9^

We have previously shown that the antibody responses elicited by 2 doses of BNT162b2 were diminished in adolescents with kidney diseases.^10^ However, data pertaining to the immunogenicity and safety of COVID-19 vaccines among the pediatric population remain extremely scarce.^11,12^ Data concerning the humoral and cellular immunogenicity to 3-dose primary series of COVID-19 vaccines among young children with kidney diseases, as well as immunogenicity against the Omicron variant, are lacking.

We initiated a 3-year non-randomized study (NCT04800133) to investigate the use of COVID-19 vaccines in children and adolescents in Hong Kong. In the present interim analysis, we recruited younger children and adolescents aged 0-18 years with advanced chronic kidney disease (CKD, stage 3 or above), and those on immunosuppressive therapy, on chronic dialysis and post-kidney transplant as they are clinically vulnerable. We evaluated the immunogenicity and safety in participants who had initiated or completed a 3-dose primary series of either CoronaVac (age <5 years), or BNT162b2 (0.1ml dose for 5-11 years, and 0.3ml dose for 11-18 years).

## METHODS

### Study Design

COVID-19 Vaccination in Adolescents and Children (COVAC; NCT04800133) is a non-randomized study investigating the safety and immunogenicity of BNT162b2 and CoronaVac, in healthy children and adolescents or those with pediatric illnesses as previously described.^10,13,14^ The study was approved by the University of Hong Kong/Hong Kong West Cluster Hospital Authority Institutional Review Board (UW21-157).

### Participants

The current analysis included children and adolescents, aged 0-18 years, with advanced chronic kidney disease (CKD, stage 3 or above), and those on immunosuppressive therapy, on chronic dialysis and post-kidney transplant. All patients received at least two doses of COVID-19 vaccine at the time of analysis. Participants with no known history of kidney diseases were excluded from this analysis.

### Procedures

Potential participants were recruited by pediatricians from the Pediatric Nephrology Centre, Hong Kong Children’s Hospital and the Department of Pediatrics and Adolescent Medicine, Queen Mary Hospital in Hong Kong. The Pediatric Nephrology Centre in Hong Kong Children’s Hospital is the territory-wide, designated pediatric referral center for complicated kidney diseases and chronic kidney replacement therapy, including dialysis and transplant. Clinicians obtained informed consent from participants aged 18 years. Informed assent was obtained from participants aged below 18, and consent was obtained from their parents or legally acceptable representatives. Participants were either unvaccinated, or partially vaccinated at the time of enrollment. Demographic information was reported by participants, and clinical information were extracted from their health records. In our study, 0.5ml CoronaVac was offered to participants aged below 5 years, 0.1ml BNT162b2 was offered to participants aged 5-11 years as the original pediatric formulation was not available in Hong Kong, and 0.3ml BNT162b2 was offered to participants aged 11-18 years. Participants who received an alternative COVID-19 vaccine as the first or second dose prior to joining the study would complete the remaining doses with the vaccine brand indicated for their age in our study protocol. Additional consent and assent were obtained for dosage switching to 0.3ml for participants who became 12 years old after initiating the primary series. CoronaVac and BNT162b2 were administered via the intramuscular route to the deltoid or the anterolateral thigh, or, in some for 0.3ml BNT162b2, by an intradermal inoculator (MicronJet600, NanoPass Technologies, Nes Ziona, Israel) to the deltoid. Doses 2 and 3 were given at least 14 and 28 days after the preceding dose. Vaccination was deferred in patients with SARS-CoV-2 infection and was resumed or initiated 28 days later. All participants were observed by a nurse or pediatrician after each dose for at least 15 minutes after vaccination on site. Blood-taking and safety data collection were performed pre-dose 1, pre-dose 2, post-dose 2 (14-42 days after dose 2), pre-dose 3 (blood-taking was only performed for participants who received dose 3 more than 56 days after their post-dose 2 visit), and post-dose 3 (14-42 days after dose 3).

### Immunogenicity

#### Humoral

Primary outcomes on humoral immunogenicity included wild-type (WT, i.e. ancestral) Spike receptor-binding domain (S-RBD) IgG ELISA and WT surrogate virus neutralization test (sVNT). While the S-RBD IgG is a binding antibody assay, the sVNT is a functional antibody assay which reflects blocking of S-RBD and human ACE2 receptor by vaccinee sera and correlates with the gold-standard plaque reduction neutralization test.^15^ Secondary outcome was Omicron BA.1 sVNT. In-house WT S-RBD IgG enzyme-linked immunosorbent assay (ELISA) were carried out as previously published.^13,16^ sVNT was conducted as per manufacturer’s instructions (GenScript Inc, Piscataway, USA).^16^

#### Cellular

Primary outcomes on cellular immunogenicity included antiviral cytokine-expressing (IFN-γ^+^ or IL-2^+^) helper (CD4^+^) or cytotoxic (CD8^+^) T cell responses against SARS-CoV-2 S (and N and M) proteins. They were examined by intracellular cytokine staining on flow cytometry after stimulation with SARS-CoV-2 15-mer peptide pool(s) (Miltenyi Biotec, Bergisch Gladbach, Germany) as described previously.^13,14,17^ SARS-CoV-2 S peptide pool was used for BNT162b2 while S, N and M peptide pools were used for CoronaVac as it is an inactivated whole-virus vaccine. Frequencies of T cell responses against S, N and M peptide pools were summated for CoronaVac. Secondary outcomes on cellular immunogenicity included T cell responses to stimulation by BA.1 S (and N and M) mutation pools, which consists of 15-mer peptides spanning the regions with BA.1-associated mutations only, were examined with WT reference pools consisting of WT sequences in the same regions as the comparator (Miltenyi Biotec, Bergisch Gladbach, Germany for S mutation pools and ChinaPeptides, Shanghai, China for N and M mutation pools).^14,17^

#### Analysis

Outcome data on immunogenicity were transformed by taking natural logarithm of the values and compared longitudinally using paired t test. Negative values, i.e., those below the limit of detection (LOD), limit of quantification (LOQ) or cut-off, were imputed as half the limit or cut-off and included in the analyses. Certain WT T cell and Omicron-specific antibody and T cell responses could only be performed in participants with sufficient blood sample volume. Additional details are available in the Supplemental Methods.

### Safety and reactogenicity

Participants reported pre-specified adverse reactions (ARs) in an online or paper-based diary for 7 days after vaccination. Unsolicited adverse events (AEs) were captured for up to 28 days after each dose. Severe AEs, including life-threatening complications, unanticipated or prolonged hospitalizations, disabilities, deaths and birth defects of their offspring, or breakthrough COVID-19, would be monitored for 3 years after vaccination. We also monitored graft rejection among kidney transplant recipients and disease flare among those with glomerular disease following vaccination. Adverse events reported were reviewed by investigators, who determined any possibility of causal relationship with the study vaccine.

## RESULTS

### Participant composition

A total of 64 children and adolescents with kidney diseases (mean age 12.2, interquartile range 9.4-16.2, and 32 male) received at least one dose of COVID-19 vaccine, including 5 aged 1-5 years for CoronaVac, 25 aged 5-11 years for 0.1ml BNT162b2, and 34 aged 11-18 years for 0.3ml BNT162b2 (Table 1). Thirteen patients were on dialysis, 15 patients underwent kidney transplant, and 31 patients were on immunosuppressives alone at the time of dose 1. All participants had Asian heritage. The details of these patients, grouped according to their treatment, are summarized in Table S1. Five patients aged 14-18 years elected to receive dose 3 BNT162b2 intradermally.

**TABLE 1.** Participant profile by vaccine type and age. eGFR (estimated glomerular filtration rate) is estimated by modified Schwartz equation for participants below age 18 years and CKD-EPI formula for those above. KRT, kidney replacement therapy

|  | All patients | CoronaVac,<br><5 years | BNT162b2,<br>5-11 years, 0.1ml | BNT162b2, 11-18<br>years, 0.3ml |
| --- | --- | --- | --- | --- |
| Number of participants *N* | 64 | 5 | 25 | 34 |
| Age mean (interquartile range) | 12.2 (9.4 – 16.2) | 3.3 (3.1-3.6) | 9.1 (8.6-12.1) | 15.7 (10.5-16.4) |
| Sex male *n*:female *n* | 32:32 | 2:3 | 12:13 | 18:16 |
| Number of participants completing 2 doses *n* (%) | 64 (100%) | 5 (100%) | 25 (100%) | 34 (100%) |
| Number of participants completing 3 doses *n* (%) | 51 (80%) | 5 (100%) | 17 (68%) | 29 (85%) |
| **Primary kidney diagnosis *n* (%)** |  |  |  |  |
| CAKUT | 10 (16%) | 1 (20%) | 4 (16%) | 5 (15%) |
| Glomerular diseases | 36 (56%) | 3 (60%) | 15 (60%) | 18 (53%) |
| Hereditary | 11 (17%) | 1 (20%) | 4 (16%) | 6 (18%) |
| Miscellaneous | 7 (11%) | 0 (0%) | 2 (8%) | 5 (15%) |
| **Treatment modality *n* (%)** |  |  |  |  |
| No KRT | 36 (56%) | 4 (80%) | 16 (64%) | 16 (47%) |
| On immunosuppression | 31 (48%) | 2 (40%) | 14 (56%) | 15 (44%) |
| **KRT** |  |  |  |  |
| Kidney transplant | 15 (23%) | 0 (0%) | 7 (28%) | 8 (24%) |
| Dialysis | 13 (20%) | 1 (20%) | 2 (8%) | 10 (29%) |
| Vintage of KRT mean (range) | 5.7 (1-18) | 2 (2) | 6.2 (2-10) | 5.6 (1-18) |
| **Concurrent immunosuppression within 3 months *n* (%)** |  |  |  |  |
| Mycophenolate mofetil | 32 (50%) | 1 (20%) | 13 (52%) | 18 (53%) |
| Prednisolone | 38 (59%) | 1 (20%) | 15 (60%) | 22 (65%) |
| Azathioprine | 3 (5%) | 0 (0%) | 0 (0%) | 3 (9%) |
| Cyclosporine A | 2 (3%) | 0 (0%) | 1 (4%) | 1 (3%) |
| Tacrolimus | 29 (45%) | 1 (20%) | 13 (52%) | 15 (44%) |
| Everolimus | 4 (6%) | 0 (0%) | 3 (12%) | 1 (3%) |
| Hydroxychloroquine | 6 (9%) | 0 (0%) | 1 (4%) | 5 (15%) |
| Rituximab | 3 (5%) | 1 (20%) | 0 (0%) | 2 (6%) |
| None | 14 (22%) | 3 (60%) | 3 (12%) | 8 (24%) |
| **Immunosuppression within 1 year *n* (%)** |  |  |  |  |
| Rituximab | 9 (14%) | 1 (20%) | 2 (8%) | 6 (18%) |
| **Kidney function and proteinuria mean (range)** |  |  |  |  |
| eGFR (ml/min/1.73m<sup>2</sup>) | 88.1 (20-234) | 97.0 (20-234) | 91.2 (20-184) | 84.0 (24-145) |
| Urine protein/creatinine ratio (mg/mmol) | 33.9 (1.6-140) | 22.7 (14-32) | 29.8 (1.6 -140) | 40.2 (6-108) |
| **Blood count and comorbidities** |  |  |  |  |
| Hypertension *n* (%) | 38 (59%) | 4 (80%) | 12 (48%) | 22 (65%) |
| Absolute lymphocyte count (x10<sup>6</sup>/ml) mean (range) | 2.5 (0.48-6.47) | 4.5 (3.27-6.47) | 2.9 (1-5.59) | 2.0 (0.48-5.18) |

### Primary immunogenicity analysis of longitudinal antibody responses by vaccine type

We studied antibody responses as they have been shown to correlate with vaccine efficacy against symptomatic COVID-19.^18^ We tracked antibody responses against WT SARS-CoV-2, including S-RBD IgG for binding antibody and sVNT for neutralization, longitudinally from the pre-vaccine baseline to post-dose 3 by vaccine type in Fig. 1A and 1B respectively. S-RBD IgG and sVNT responses rose significantly for all 3 vaccine types/ages after 3 doses when compared to pre-dose 1 on paired t test after natural logarithmic transformation. For patients who received the 3-dose primary series of CoronaVac (aged <5 years), all children tested positive for S-RBD IgG (above LOD of 0.5, considered ‘seropositive’) and had a high geometric mean sVNT% level of 94.0%. For patients who received the 3-dose primary series of BNT162b2, 73.9% and 93.8% of the participants aged 5-11 years (who received 0.1ml BNT162b2) were seropositive after doses 2 and 3, with geometric mean sVNT% levels of 51.7% and 78.6%. These figures were similar to 11-18 years age group (who received 0.3ml BNT162b2) where 81.8% and 90.9% of the participants were seropositive, with geometric mean sVNT% levels of 60.5% and 80.0% after doses 2 and 3 respectively. Compared against 2 doses only, an additional third dose of 0.1ml and 0.3ml BNT162b2 both elicited significant increases in S-RBD IgG and sVNT. Four patients who received 2 intramuscular and 1 intradermal dose of 0.3ml BNT162b2 had a high geometric mean sVNT% level of 95.7% post-dose 3. S-RBD IgG and sVNT responses were also presented by kidney replacement therapy status in Table 2.

**TABLE 2.** Antibody responses including S-RBD IgG and surrogate virus neutralization test (sVNT) against wild-type SARS-CoV-2 by treatment status. Number of seropositive patients by S-RBD IgG were given by n/N (%) and geometric mean sVNT with 95% confidence intervals (CIs) were shown. KRT, kidney replacement therapy

|  | No KRT |  |  | On KRT |  |
| --- | --- | --- | --- | --- | --- |
|  | No IS | Any IS | Rituximab | Dialysis | Transplant |
| **S-RBD IgG seropositivity (%)** |  |  |  |  |  |
| **Pre-dose 2** | 4/4 (100%) | 14/29 (48.3%) | 2/9 (22.2%) | 8/12 (66.7%) | 3/12 (25.0%) |
| **Post-dose 2** | 5/5 (100%) | 23/30 (76.7%) | 5/9 (55.6%) | 11/11 (100.0%) | 10/15 (66.7%) |
| **Post-dose 3** | 4/4 (100%) | 19/20 (95.0%) | 3/4 (75.0%) | 8/8 (100.0%) | 9/11 (81.8%) |
| **Geometric mean sVNT% level (95% CI)** |  |  |  |  |  |
| **Pre-dose 2** | 67.0% (47.3-100.6%) | 28.6% (21.7-37.6%) | 21.3% (12.5-36.4%) | 39.4% (24.6-63.0%) | 21.4% (14.1-32.4%) |
| **Post-dose 2** | 96.5% (95.0-97.9%) | 57.2% (42.8-76.4%) | 30.7% (15.6-60.6%) | 91.5% (85.5%-97.9%) | 37.0% (22.6- 60.6%) |
| **Post-dose 3** | 97.4% (96.1-98.8%) | 86.5% (71.1-105.2%) | 60.8% (13.8-267.9%) | 95.9% (93.9-98.0%) | 59.4% (35.9- 98.2%) |

**FIG. 1.**
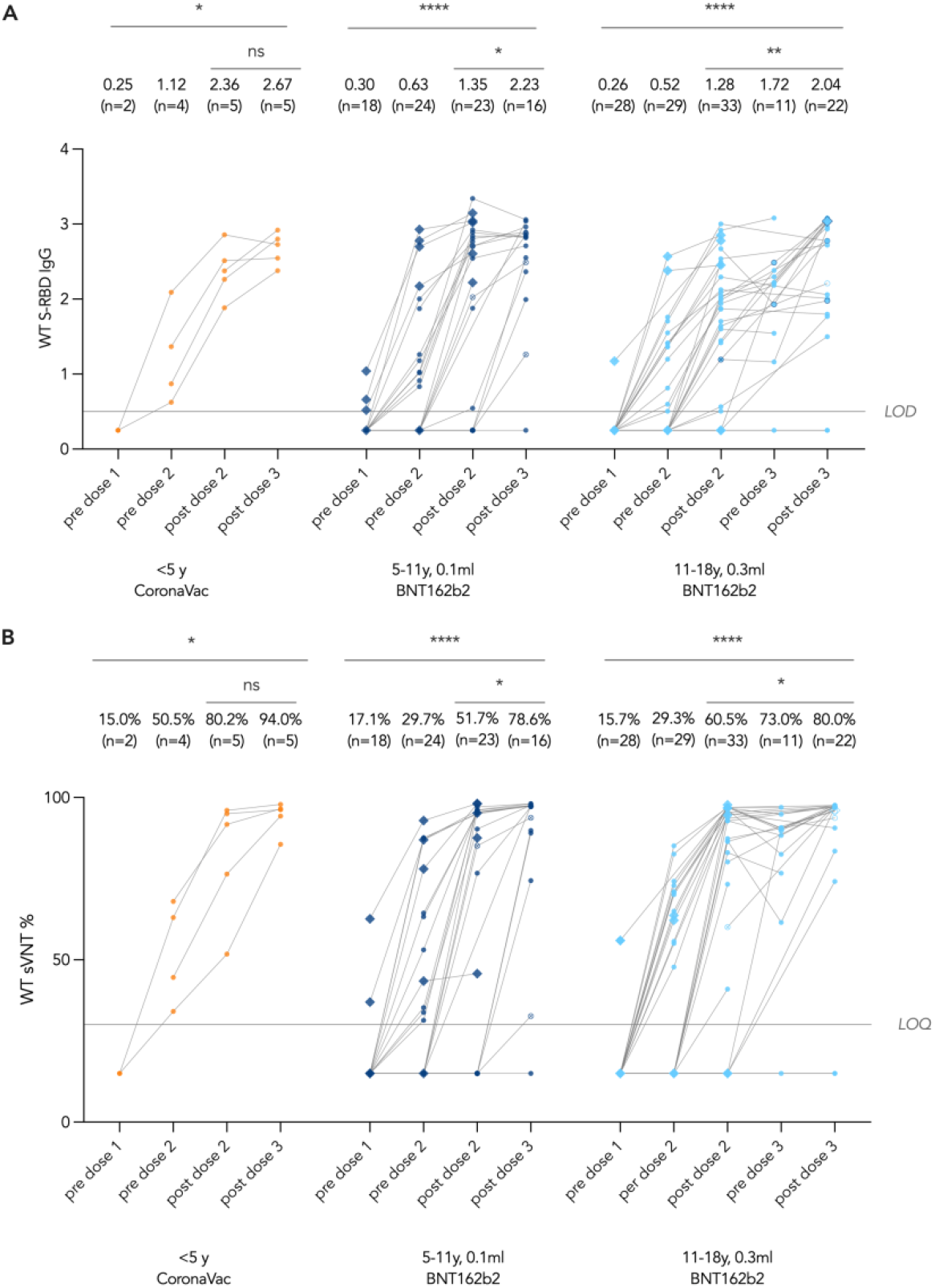
Antibody responses against wild-type SARS-CoV-2 including (A) S-RBD IgG for binding and (B) surrogate virus neutralization test (sVNT) for neutralization. Matched pre-dose 1 or post-dose 2 tests were compared with post-dose 3 by paired t test after natural logarithmic transformation, and p values are denoted by asterisks (*, P<0.05; **, P<0.01; ****,P<.0001; ns, not significant). Geometric means (GM) are shown with center lines and stated above each column. Limits of detection and quantification (LOD and LOQ) were drawn as grey lines. Data points of patients who received intradermal 0.3ml BNT162b2 as dose 3 were depicted as rhombi, and those of infected patients were drawn to be hollow beginning from after the infection.

### Primary immunogenicity analysis of longitudinal T cell responses by vaccine type

We also studied T cell responses as they protect against progression to severe COVID-19.^19^ For patients who received CoronaVac (< 5 years), IFN-γ^+^ antiviral CD4^+^ helper and CD8^+^ cytotoxic T cell responses against SARS-CoV-2 S, N and M proteins were examined longitudinally in Fig 2. In addition, CD4^+^ and CD8^+^ T cells expressing another antiviral cytokine, IL-2, were also studied (Fig. S1A-D). In patients who received CoronaVac (<5 years), we detected a significant increase of SNM-specific IFN-γ^+^ CD4^+^ T cell response (Fig. 2B). Other T cell responses also trended to increase, albeit insignificantly. Yet, all 5 patients who received 3 doses of CoronaVac were positive for either S-specific or SNM-specific IFN-γ^+^ CD4^+^ and CD8^+^ T cells.

**FIG. 2.**
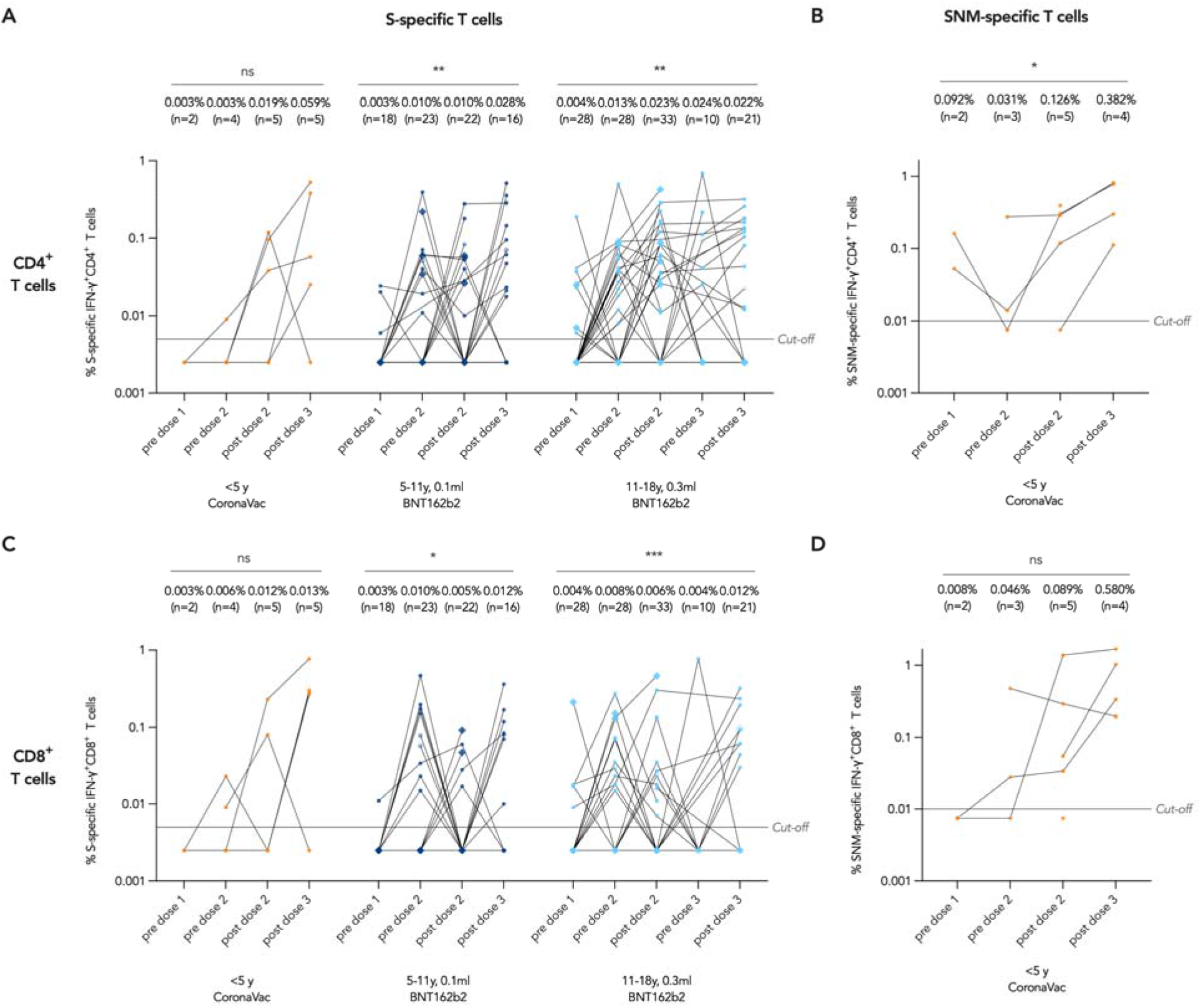
IFN- γ^+^ T cell responses against wild-type SARS-CoV-2 proteins. T cell responses against S protein were tested for all participants while responses against S, N and M proteins were only tested for CoronaVac. Matched pre-dose 1 and post-dose 3 tests were compared by paired t test after natural logarithmic transformation, and p values are denoted by asterisks (*, P<.05; **, P<0.01; ***, P<0.001; ns, not significant). Geometric means (GM) are shown with center lines and stated above each column. Cut-offs were drawn as grey lines. Data points of patients who received intradermal 0.3ml BNT162b2 as dose 3 were depicted as rhombi, and those of infected patients were drawn to be hollow beginning from after the infection.

For patients who received BNT162b2, T cell response against S protein only was examined. After 3 doses of BNT162b2, we detected significant increases in all S-specific IFN-γ^+^ and IL-2^+^ CD4^+^ and CD8^+^ T cell responses (Fig. 2A, 2C), except for S-specific IL-2^+^ CD8^+^ T cell response in those aged 5-11 years who received 0.1ml BNT162b2 (Fig. S1). Four patients who received an intradermal dose 3 of 0.3ml BNT162b2 appeared to have a similar level of T cell responses as those who received intramuscular dose 3.

### Antibody and T cell responses against Omicron BA.1

As a secondary outcome, Omicron BA.1 neutralization was evaluated by sVNT. When compared with paired WT sVNT, we found a significant lower sVNT% level against Omicron BA.1 following dose 2 (64.1% vs 18.5%, P<0.0001) and dose 3 (84.0% vs 27.2%, P<0.0001) (Fig. 3A), indicating that BA.1 markedly evades neutralization compared to WT in kidney patients. However, we observed a significant increase in geometric mean BA.1 sVNT% level with dose 3 (post-dose 2 vs post-dose 3, 18.5% vs 27.2%, P=0.006).

**FIG. 3.**
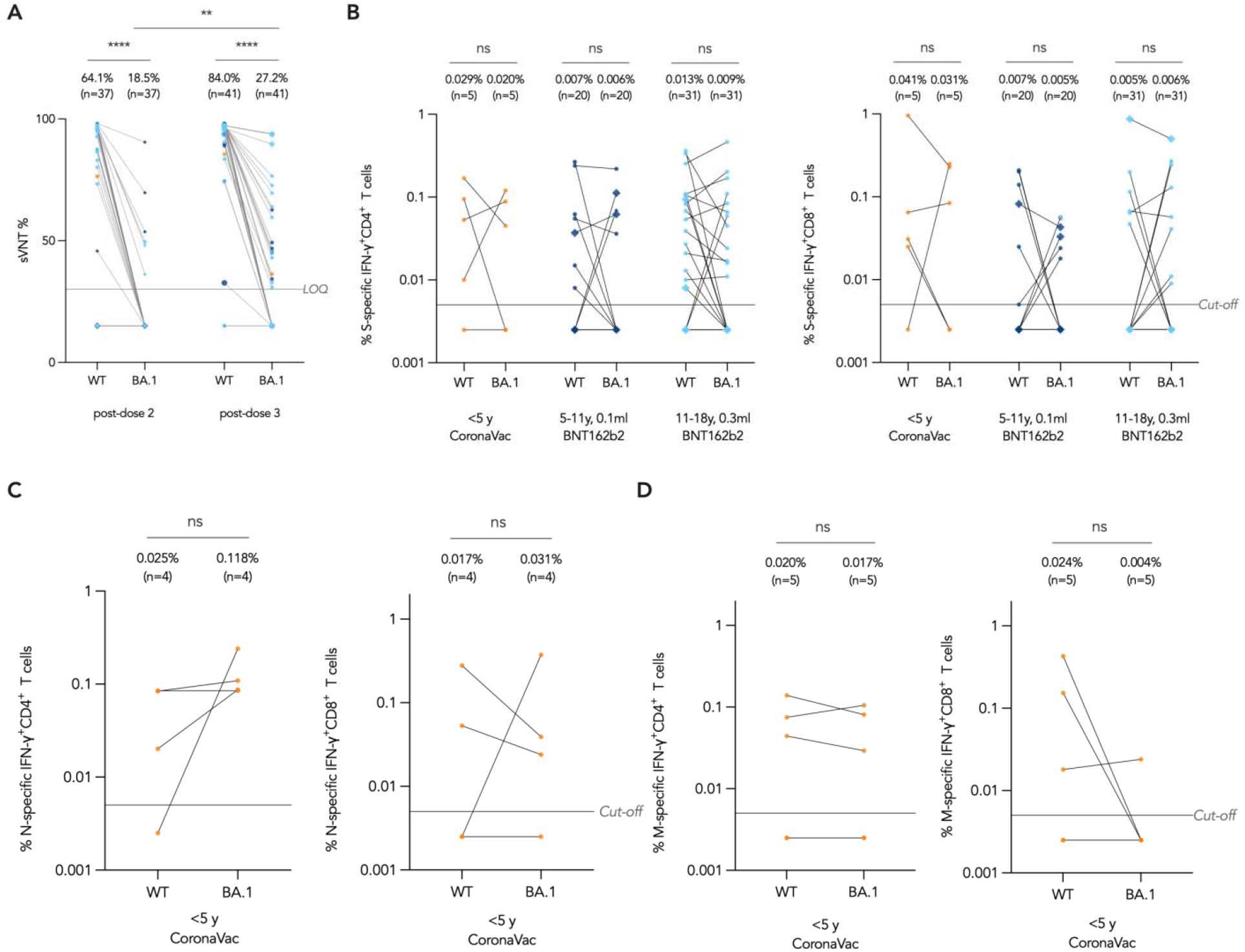
Antibody and T cell response against Omicron BA.1. Matched WT and BA.1 tests were compared by paired t test after natural logarithmic transformation, and the p values are denoted by asterisks (**, P<0.01; ****, P<0.0001; ns, not significant). Geometric means (GM) are shown with center lines and stated above each column. Limit of quantification (LOQ) and cut-offs were drawn as grey lines. Data points were colored orange, navy blue and cyan according to vaccine type and age, referring to <5 years CoronaVac, 5-11 years 0.1ml BNT162b2 and 11-18 years 0.3ml BNT162b2 respectively. Data points of patients who received intradermal 0.3ml BNT162b2 as dose 3 were depicted as rhombi, and those of infected patients were drawn to be hollow beginning from after the infection.

We then assayed IFN-γ^+^ CD4^+^ and CD8^+^ T cells responses against Omicron BA.1 mutations in S, N and M after 2 doses of vaccine. Notably, T cell responses against Omicron BA.1 and WT S protein were comparable after 2 doses of any vaccine (Fig. 3B). Similarly, in patients aged <5 years who received CoronaVac, N-specific and M-specific T cell responses were comparable between Omicron BA.1 and WT (Fig. 3C-D), suggesting T cell responses were not diminished by mutations in Omicron BA.1.

### Correlation of immunogenicity with vaccine type and treatment modality

We explored correlation of immunogenicity with age or type of vaccine, and treatment as a secondary analysis. We focused on post-dose 2 WT sVNT because antibody assays are more sensitive than T cell assays and correlates with plaque neutralization better than S-RBD IgG. No correlation with immunogenicity was found for age or type of vaccine. However, compared to non-dialysis patients who were not on immunosuppressives, a significantly lower sVNT level was detected for both kidney transplant recipients (geometric mean ratio of sVNT% 0.38, 95% CI 0.16-0.90; P=0.03) and patients who received rituximab within 1 year preceding vaccination (geometric mean ratio of sVNT% 0.32, 95% CI 0.13-0.77; P=0.02) (Fig. 4). In contrast, patients on chronic dialysis demonstrated a similar antibody response to non-dialysis patients not on immunosuppression (P=0.27).

**FIG. 4.**
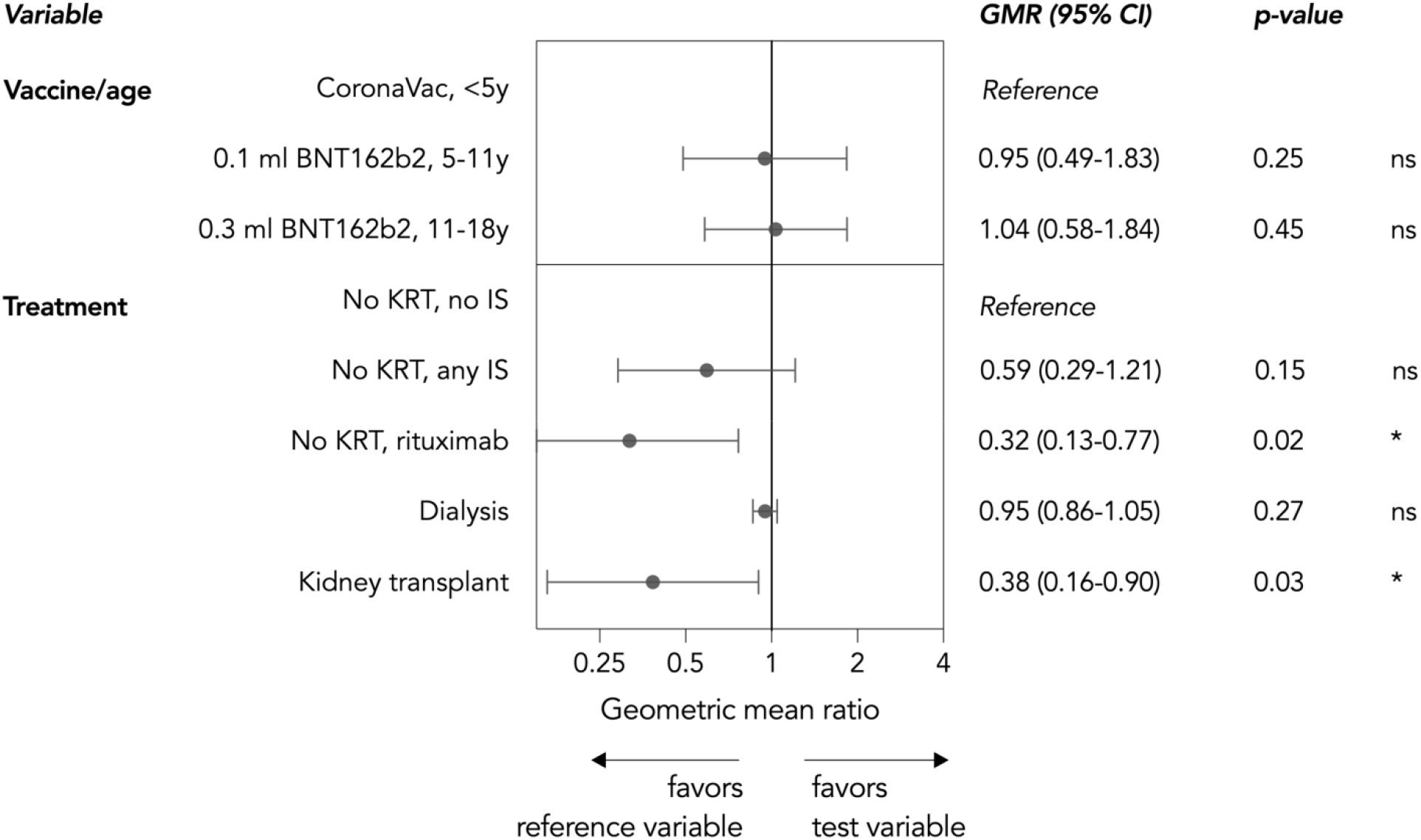
Correlation of surrogate virus neutralization test (sVNT) % levels with vaccine type (or age) and treatment. sVNT results at 1 month after 2 doses were compared by unpaired t test after natural logarithmic transformation. KRT, kidney replacement therapy; IS, immunosuppressives

### Breakthrough COVID-19 cases and hybrid immunity

From January 2022, Hong Kong was hit by the first major wave of COVID-19 resulting from Omicron BA.2 variant. Eleven patients (17% of all participants) reported COVID-19 during that time, including 7 (11%) before and 4 (6%) after initiating COVID-19 vaccination. All patients reported mild COVID-19 without the need of hospitalization. No mortality was reported. Two patients had a longer clinical course with rapid antigen test positivity for 14 and 17 days. They were both unvaccinated and treated with immunosuppressive therapy for underlying glomerular disease. Two patients had breakthrough COVID-19 after completing 3 doses, and both had a high WT sVNT% level above 90% (93.7% and 90.6%) tested shortly before breakthrough infection. Both patients were managed conservatively without complications.

We also investigated the effect of hybrid immunity, which refers to the synergized immune response from both vaccination and infection, in patients who were infected prior to vaccination (Fig. S2A-C). Before vaccination, only two patients out of 7 seroconverted to infection alone, and one of them had a positive yet weak neutralizing response (Fig. S2A). After receiving 2 doses of vaccine, infected patients showed significantly boosted S-RBD IgG and WT sVNT responses (Fig. S2A). When compared to uninfected patients who received 2 doses of vaccine, infected and vaccinated patients also had a higher Omicron BA.1 sVNT (Fig. S2B). For T cell responses among infected patients, S-specific IFN-γ^+^ CD4^+^ T cells were significantly increased after 2 doses compared to pre-dose 1, and S-specific IFN-γ^+^ CD8^+^ T cells showed an upward trend as well (Fig. S2C).

### Safety and reactogenicity

We tracked ARs for 7 days and AEs for 28 days after each dose. COVID-19 vaccination appeared tolerable in our patients. Patients receiving either vaccine brand mainly reported mild and moderate ARs (Fig. 5). Two and 5 AEs were reported within 28 days after 0.1ml and 0.3ml BNT162b2, including 2 reports after dose 1 and 5 reports after dose 2. They included chest discomfort (n=1), palpitations (n=2), tachycardia (n=1), epistaxis (n=1), insomnia (n=1), and tinnitus (n=1). All AEs were grade 1 except for the report of tinnitus, which was grade 2. None of the kidney transplant recipients developed graft rejection during the follow-up period. Of the 31 non-dialysis patients with glomerular disease, 2 patients (6.4%) developed disease relapse closely following vaccination, 1 of whom required hospitalization. Two more patients experienced relapse, but their relapses could be attributable to acute COVID-19 and to B cell reconstitution 12 months after rituximab, respectively. Four patients reported a non-fatal resolved severe AE after study vaccine administration, including one for nephrotic syndrome relapse, one with suspected myopericarditis, one with prolonged post-vaccine fever and one with catheter-associated infection. All patients with an SAE recovered without complications. More details regarding adverse events are available in Supplemental Information.

**FIG. 5.**
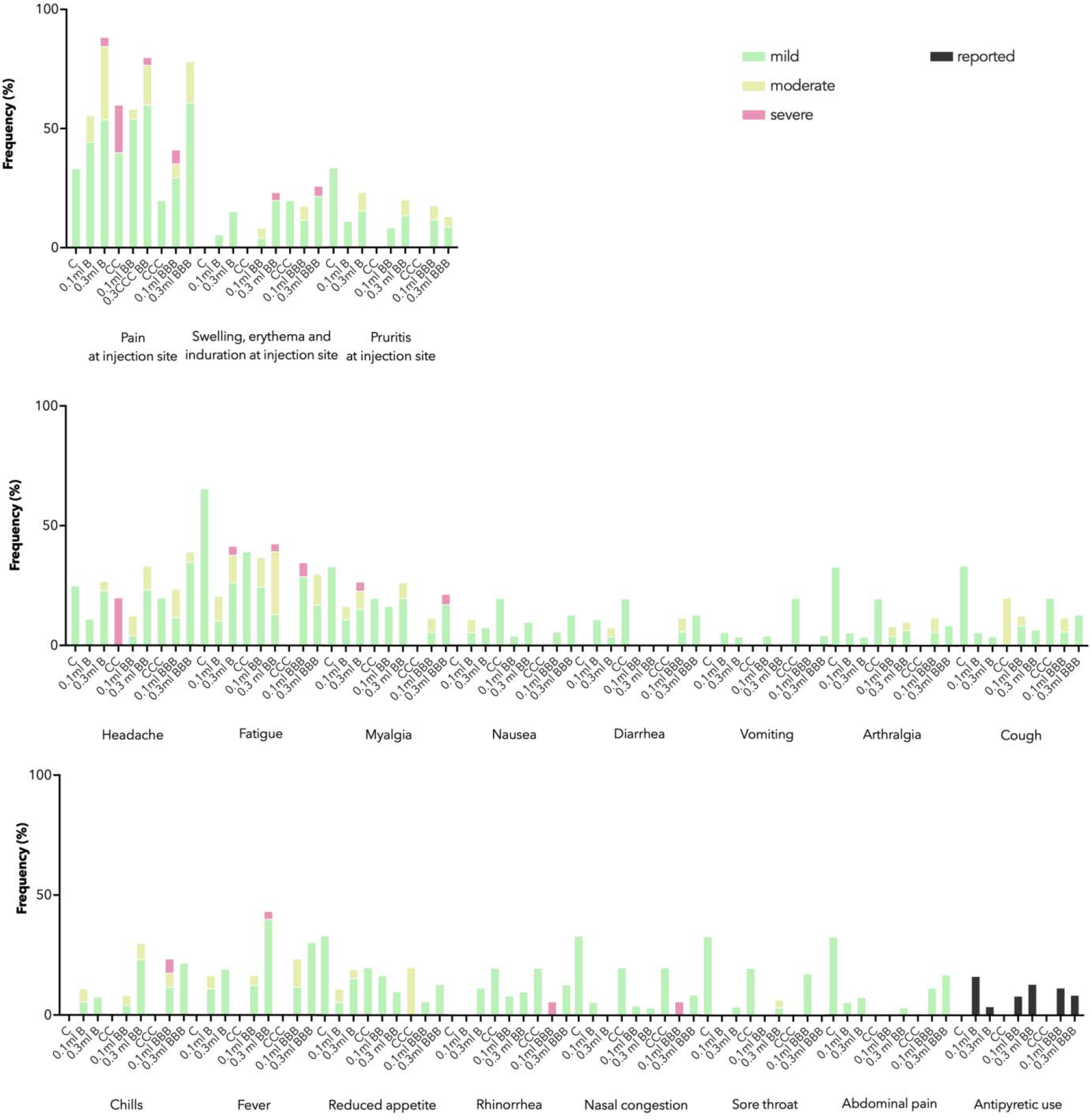
Adverse reactions (ARs) and antipyretic use reported 7 days after each dose by vaccine type. Stacked bar chart shows ARs by maximal severity in different colors. C - CoronVac, 0.1ml or 0.3ml B - BNT162b2.

## DISCUSSION

In this study, we demonstrated that an accelerated, 3-dose COVID-19 vaccine schedule with CoronaVac (<5 years) and BNT162b2 at age-appropriate dose (5-18 years) elicited favorable antibody and T cell responses. However, weaker antibody responses were found in patients who received kidney transplant or rituximab. Although Omicron BA.1 mutations led to partial neutralization escape, dose 3 significantly enhanced neutralization capacity. Importantly, T cell responses against Omicron BA.1 were comparable to that with WT, suggesting effective protection from severe COVID-19 by vaccination.

We showed a significant induction of S-RBD IgG and WT sVNT antibody responses regardless of vaccine type and age. Antibody response is the first line of defense against infection and has been shown to correlate with efficacy against COVID-19,^18^ Therefore, our results suggest that 3 doses of COVID-19 vaccines could offer protection against COVID-19 in children and adolescents with kidney diseases. In contrast to healthy adolescents who have a 100% seropositivity after a single dose of BNT162b2,^13^ 4 out of 47 (9%) patients failed to develop antibody responses after 3 doses of vaccine. Similarly, studies on adult patients who are on dialysis or post-kidney transplant also demonstrated weak antibody responses, related to the immunosuppressed state in kidney failure and immunosuppressive therapy.^20,21^ In our study, kidney transplant recipients and patients receiving rituximab for glomerular disease were identified to have lower antibody response. This is likely to be due to intense humoral immunosuppression by triple therapy post-transplant or by rituximab. Rituximab has emerged as an important therapeutic option in several pediatric immune-mediated glomerular diseases, such as idiopathic nephrotic syndrome and ANCA-associated vasculitis glomerulonephritis.^2,22,23^ Our data showed that rituximab exposure up to 1 year prior to vaccination resulted in poor immunogenicity. Furthermore, patients with glomerular disease are at risk of relapse and often require multiple courses of rituximab to maintain disease remission.^22^ Therefore, the timing of vaccination and rituximab should be individualized, balancing the risk of infection and disease relapse. Fortunately, our current findings did not suggest an overtly increased risk of graft rejection and disease relapse following vaccination, indicating the safety of COVID-19 vaccination in children and adolescents with kidney diseases.

The SARS-CoV-2 Omicron variant has caused repeated surges in COVID-19 cases and hospitalizations across the globe, with more than 30 mutations in S protein which cause extensive neutralization escape and reduce the effectiveness of COVID-19 vaccines.^24-26^ According to our data, when compared with WT sVNT, we found that a significantly lower sVNT% level against Omicron BA.1 following doses 2 and 3, signifying partial neutralization escape. However, neutralization capacity was significantly enhanced by dose 3 from 18.5% to 27.2%. We postulate that further booster doses may further optimize the immunogenicity and protection in children and adolescents with kidney diseases, as a fourth dose has been found to seroconvert adult kidney transplant recipients who were seronegative after 3 doses.^27^ Interestingly, our data suggested that T cell responses remained robust against Omicron even with a WT vaccine, in line with observations in healthy adolescents.^14,17^ T cell responses against SARS-CoV-2, including towards non-S proteins,^28^ may protect against progression to severe COVID-19 after a breakthrough infection,^19^ even against variants of concerns with major neutralization escape.

We only observed mild infections after vaccination in our cohort. It was known that either a post-vaccine breakthrough infection or vaccination following infection improves the magnitude and diversity of immune response, leading to hybrid immunity that could be longer-lasting and cross-reactive towards novel variants.^29^ Our data showed that only 28% patients seroconverted to infection alone. Yet, Omicron BA.1 sVNT after 2 doses among infected patients was significantly higher than that of uninfected vaccinated patients. This further supports the need of vaccination in pediatric kidney patients even after SARS-CoV-2 infection, as this will generate hybrid immunity, and enhance protection against future variants. Although few of our vaccinated participants experienced a breakthrough infection, allow us to investigate their immune responses, those vaccinated and experienced a breakthrough infection should likewise complete all recommended doses for maximal protection. Nonetheless, we advocate for continued personal hygiene measures to minimize the chance of a severe breakthrough COVID-19 in children and adolescents with kidney diseases.

In this study, we vaccinated young children under age 5 with full-dose CoronaVac, an inactivated COVID-19 vaccine, and children aged 5-11 years with 0.1ml BNT162b2. Real-world effectiveness data in Hong Kong collected during the BA.2.2 wave showed comparable effectiveness against severe and fatal COVID-19 after 3 doses of either vaccine brand in adults,^26^ and against infection after 2 doses of either vaccine brand in children and adolescents.^8^ The investigators preferred BNT162b2 for children and adolescents with kidney diseases as there were more effectiveness data on mRNA COVID-19 vaccines in immunocompromised patients.^30,31^ Yet, the pediatric formulation of BNT162b2 was not made available to Hong Kong, and we adopted dosage reduction, injecting 0.1ml for an equivalent potency of 10mcg, with reference to United Kingdom Joint Committee on Vaccination and Immunization advice for children in a clinically risk group in December 2021.^32^ In children below 5 years, as safety and immunogenicity data were published for CoronaVac and not for BNT162b2 at the time of the Omicron BA.2.2 wave in Hong Kong,^5^ we offered CoronaVac to young children and infants with kidney diseases urgently. Our findings suggest both 0.1ml BNT162b2 and CoronaVac were safe and immunogenic and support their use in children with kidney diseases.

Furthermore, due to the rapid Omicron BA.2 wave in Hong Kong during study period, we administered the vaccines using an accelerated regimen, with 14-day interval between doses 1 and 2, and 28-day interval between doses 2 and 3. While longer prime-boost intervals are associated with better immunogenicity in general,^33^ seronegativity after just 1 or 2 doses was common in our study, and it was important for our vulnerable patients to immediately attain a higher level of protection within a short timeframe before catching Omicron. We showed that the accelerated regimen was still able to elicit antibody and T cell responses significantly in children and adolescents with kidney diseases and speculate that their immune responses will also be boosted by breakthrough infections and booster doses.

Moreover, while our study conducted antibody and T cell testing under a research protocol, clinicians might be tempted to base their decisions of vaccination or boosting on SARS-CoV-2 antibody testing. However, the 2 participants who received all 3 doses of vaccines prior to infection both showed high WT sVNT antibody response (>90%) prior to acute COVID-19. This suggests antibody testing in an individual patient cannot be used to predict susceptibility to infection nor to guide booster decision.

Our study had strengths and limitations. We tracked antibody and T cell responses longitudinally in a relatively large cohort of vaccinated children and adolescents with different kidney diseases and treatments aged 1-18 years and provided the first Omicron-specific neutralization and T cell responses in this patient population. We could not assess clinical effectiveness, nor compare immunogenicity of vaccine brands in the same age groups. Sample number available for a particular test and timepoint was limited by the blood volume collected and completion of study visit by the participant. As we used 0.1ml dose of the adult formulation for BNT162b2, reactogenicity and immunogenicity could potentially differ from that of the actual pediatric formulation.

In conclusion, our findings support that an accelerated, 3-dose primary series of mRNA or inactivated COVID-19 vaccines is safe and immunogenic for children and adolescents with kidney diseases, especially among kidney transplant recipients and patients receiving rituximab therapy. Although neutralization response against Omicron was reduced, T cell response was preserved and may offer protection to children and adolescents with kidney diseases from severe COVID-19. Further studies are needed to shed light on the immunogenicity of a fourth dose and hybrid immunity in immunocompromised children in the future.

## Supporting information

Supplemental Methods and Supplemental Information: Safety and Reactogenicity

## Data Availability

All data produced in the present study are available upon reasonable request to the authors

## STATEMENT OF CONTRIBUTION

Y.L. Lau conceptualized the study. Y.L. Lau, M. Peiris, W. Tu, D. Leung, J.S. Rosa Duque, and X. Mu designed the study. Y.L. Lau led the acquisition of funding. Y.L. Lau, W. Tu, and M. Peiris supervised the project. S.M. Chan, D. Leung, X. Mu, S.M.S. Cheng, I.Y.S. Tam, and J.H.Y. Lam led the study administrative procedures. A.L.T. Ma, Y.L. Lau, J.S. Rosa Duque, E.Y.H. Chan, S. Chim, F.T.W. Ho, P.C. Tong, W.M. Lai, and M.H.L. Lee provided study-related clinical assessments and follow-up. D. Leung, S.M. Chan, S.T.K. Sze, J.H.Y. Lam, J.S. Rosa Duque, and Y.L. Lau collected clinical safety data. S.M.S. Cheng, L.C.H. Tsang, K.K.H. Kwan, and M. Peiris developed and performed S-RBD IgG and sVNT. X. Mu, Y. Chung, H.H.W. Wong, A.M.T. Lee, W.Y. Li, and W. Tu developed and performed the T cell assays. D. Leung, D.H.L. Lee, and J.H.Y. Lam curated, analyzed, and visualized the data. D. Leung, X. Mu, S.M.S. Cheng, J.S. Rosa Duque, D.H.L. Lee, J.H.Y. Lam, and S.M. Chan validated the data. D. Leung and E.Y.H. Chan drafted the manuscript and were supervised by A.L.T. Ma, J.S. Rosa Duque, and Y.L. Lau, with input from X. Mu, and S.M.S. Cheng. All authors reviewed and approved the final manuscript.

## ACKNOWLEDGEMENTS

We thank the staff at Community Vaccination Centers at Ap Lei Chau Sports Centre, Gleneagles Hospital Hong Kong, and Sun Yat-Sen Memorial Park Sports Centre. The investigators are grateful to all clinical research team members and laboratory staff of Department of Pediatrics and Adolescent Medicine of the University of Hong Kong, for their research support. We are most thankful to the study participants, as well as clinicians and laboratory staff who have provided clinical care and testing to the patients.

## DATA SHARING

All requests to share the pseudonymized data underlying the conclusions of this paper from researchers will be facilitated by the authors, subject to ethics approval. Enquiries should be addressed to.

## SUPPLEMENTAL MATERIALS

Supplemental Methods

Supplemental Information: Safety and Reactogenicity

---

**TABLE S1.** Participant profile by treatment. eGFR (estimated glomerular filtration rate) is estimated by modified Schwartz equation for participants below age 18 years and CKD-EPI formula for those above. KRT, kidney replacement therapy; NA - not applicable

|  | No KRT |  |  | On KRT |  |
| --- | --- | --- | --- | --- | --- |
| Treatment | No IS | Any IS | Rituximab | Dialysis | Transplant |
| Participants *N* | 5 | 31 | 9 | 13 | 15 |
| Age *mean (interquartile range)* | 6.5 (7.1-11.9) | 11.9 (9.6-15.9) | 13.4 (9.6-15.9) | 14.3 (9.4-16.2) | 12.9 (10.4-16.0) |
| Sex *male n:female n* | 4:1 | 15:16 | 3:6 | 4:9 | 9:6 |
| **Vaccine and age** |  |  |  |  |  |
| CoronaVac, <5 years *n (%)* | 2 (40%) | 2 (6%) | 1 (11%) | 1 (8%) | 0 (0%) |
| BNT162b2, 5-11 years, 0.1ml *n (%)* | 2 (40%) | 14 (45%) | 2 (22%) | 2 (15%) | 7 (47%) |
| BNT162b2, 11-18 years, 0.3ml *n (%)* | 1 (20%) | 15 (48%) | 6 (67%) | 10 (77%) | 8 (53%) |
| Completed 2 doses *n (%)* | 5 (100%) | 31 (100%) | 9 (100%) | 13 (100%) | 15 (100%) |
| Completed 3 doses *n (%)* | 4 (80%) | 24 (77%) | 5 (56%) | 11 (85%) | 12 (80%) |
| **Type of kidney disease *n (%)*** |  |  |  |  |  |
| CAKUT | 3 (60%) | 0 (0%) | 0 (0%) | 1 (8%) | 6 (40%) |
| Glomerular diseases | 1 (20%) | 30 (97%) | 9 (100%) | 3 (23%) | 2 (13%) |
| Hereditary | 0 (0%) | 1 (3%) | 0 (0%) | 6 (46%) | 4 (27%) |
| Miscellaneous | 1 (20%) | 0 (0%) | 0 (0%) | 3 (23%) | 3 (20%) |
| **Current immunosuppression within 3 months *n (%)*** |  |  |  |  |  |
| Mycophenolate mofetil | 0 (0%) | 21 (68%) | 7 (78%) | 1 (8%) | 10 (67%) |
| Prednisolone | 0 (0%) | 19 (61%) | 4 (45%) | 4 (31%) | 15 (100%) |
| Azathioprine | 0 (0%) | 1 (3%) | 0 (0%) | 0 (0%) | 2 (13%) |
| Cyclosporine A | 0 (0%) | 1 (3%) | 0 (0%) | 0 (0%) | 1 (7%) |
| Tacrolimus | 0 (0%) | 14 (45%) | 6 (67%) | 1 (8%) | 14 (93%) |
| Everolimus | 0 (0%) | 0 (0%) | 0 (0%) | 0 (0%) | 4 (27%) |
| Hydroxychloroquine | 0 (0%) | 5 (16%) | 1 (11%) | 1 (8%) | 0 (0%) |
| Rituximab | 0 (0%) | 3 (10%) | 3 (33%) | 0 (0%) | 0 (0%) |
| None | 5 (100%) | 0 (0%) | 0 (0%) | 9 (69%) | 0 (0%) |
| **Immunosuppression within 1 year *n (%)*** |  |  |  |  |  |
| Rituximab | 0 (0%) | 9 (29%) | 9 (100%) | 0 (0%) | 0 (0%) |
| **Kidney function *mean (range)*** |  |  |  |  |  |
| eGFR (ml/min/1.73m<sup>2</sup>) | 38.2 (20-88) | 109.7 (25-234) | 104.4 (25-234) | NA | 62.8 (24-103) |
| Urine protein/ creatinine ratio (mg/mmol) | 33.2 (1.6-91) | 37.2 (6-140) | 33.3 (6-108) | NA | 26.4 (7-74) |
| Kidney failure vintage (year) | 0 (0) | 0 (0) | 0 (0) | 3.6 (1-14) | 7.4 (3-18) |
| **Blood count and comorbidities** |  |  |  |  |  |
| Hypertension *n (%)* | 4 (80%) | 15 (48%) | 4 (44%) | 11 (85%) | 8 (53%) |
| Absolute lymphocyte count (x10<sup>6</sup>/ml) *mean (range)* | 3.9 (1.58-6.47) | 2.5 (0.48-5.22) | 2.4 (1.08-4.49) | 2.1 (0.89-5.18) | 2.5 (0.61-5.59) |

**FIG. S1.**
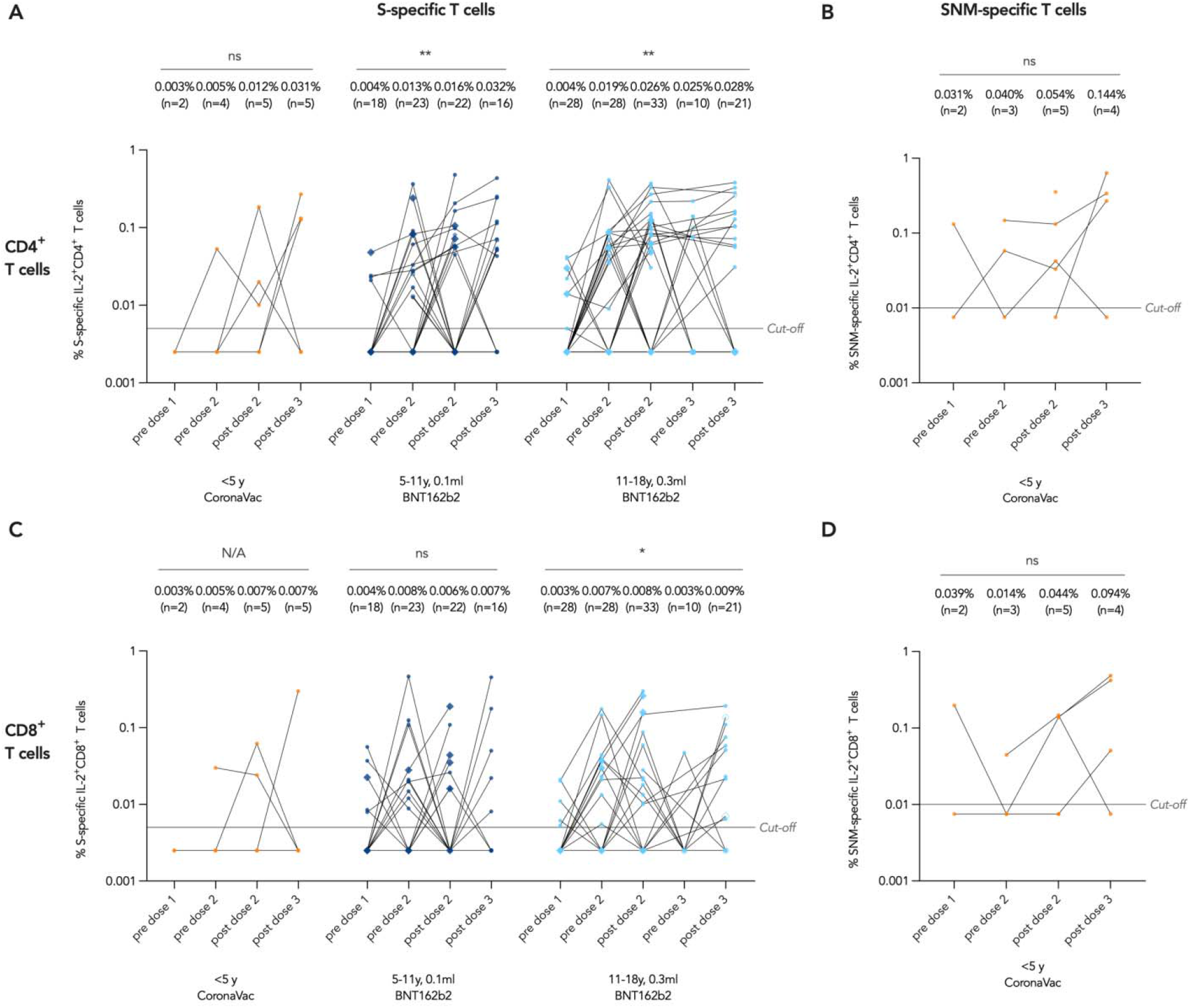
IL-2^+^ T cell responses against wild-type SARS-CoV-2 proteins. T cell responses against S protein were tested for all participants while responses against S, N and M proteins were only tested for CoronaVac. Geometric means (GM) are shown with center lines and stated above each column. Limits of detection and quantification (LOD and LOQ) and cut-offs were drawn as grey lines. Data points of patients who received intradermal 0.3ml BNT162b2 as dose 3 were depicted as rhombi, and those of infected patients were drawn to be hollow beginning from after the infection. Data from the same participant were analyzed longitudinally by paired test after natural logarithmic transformation, and the P values are denoted by asterisks (*, P<.05; **, P<0.01; ns, not significant).

**FIG. S2.**
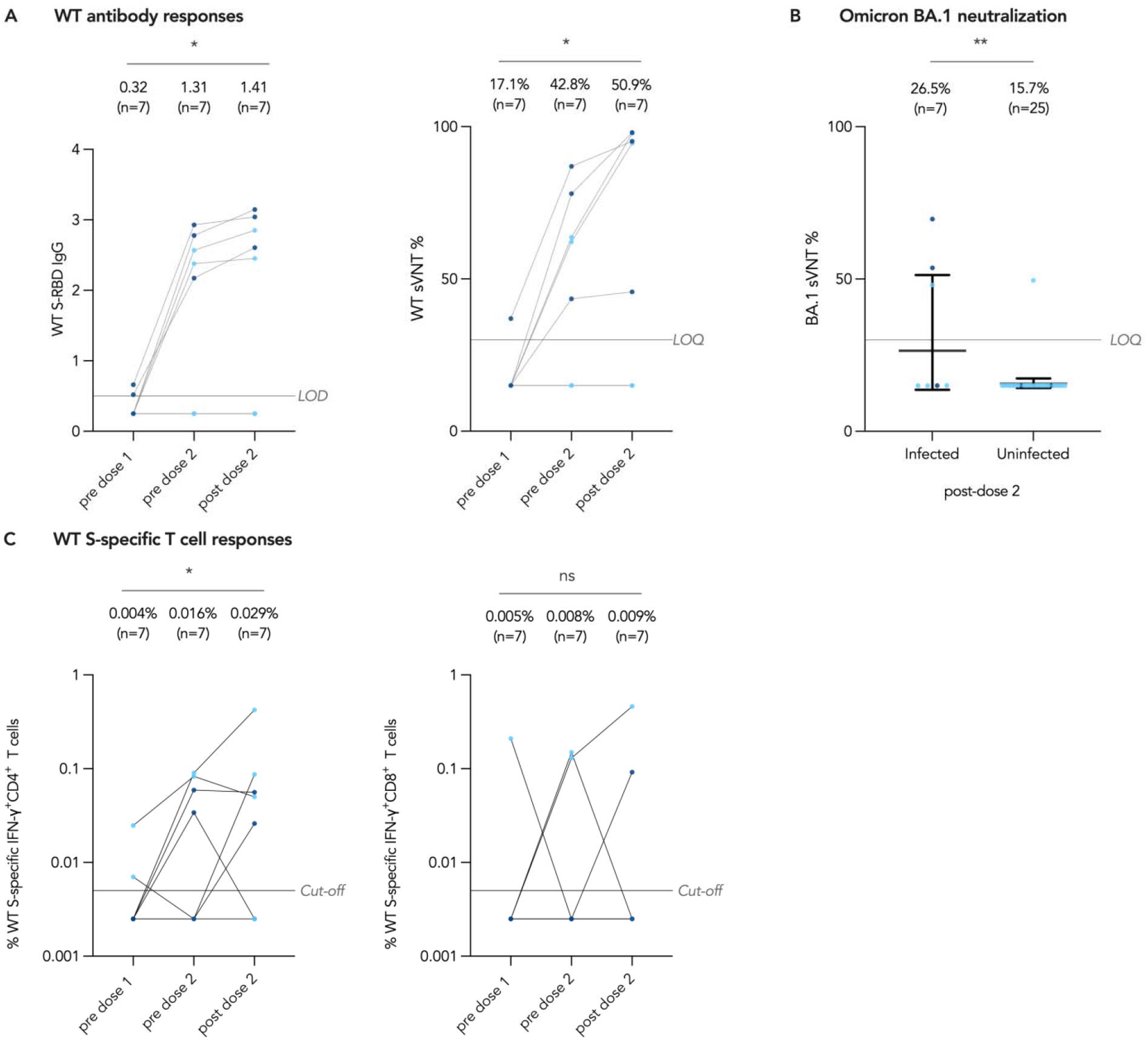
Hybrid sVNT and IFN- γ^+^ T cell responses in patients infected prior to vaccination. Geometric means (GM) are shown with center lines and stated above each column. Geometric means (GM) are shown with center lines and stated above each column. Limits of detection and quantification (LOD and LOQ) and cut-offs were drawn as grey lines. Data from the same participant were analyzed longitudinally by paired t test after natural logarithmic transformation, and the P values are denoted by asterisks (*, P<0.05; **, P<0.01; ns, not significant).

