## Supplemental Methods and Supplemental Information: Safety and Reactogenicity for "Humoral and Cellular Immunogenicity and Safety of 3 Doses of CoronaVac and BNT162b2 in Young Children and Adolescents with Kidney Diseases"

**Running Title: CoronaVac and BNT162b2 in pediatric kidney patients**

**Supplemental Materials**

Supplemental Methods	2

Supplemental Information: Safety and Reactogenicity	5

References	6

**SUPPLEMENTAL METHODS**

*SARS-CoV-2 S receptor-binding domain IgG*

Sera from participants were frozen at -80° C until testing. The S-RBD IgG enzyme-linked immunosorbent assay (ELISA) was carried out as previously described and validated.^1^ All sera were heat-inactivated at 56° C for 30 minutes before testing. In brief, S-RBD IgG ELISA plates were coated overnight with 100 ng/well of purified recombinant S-RBD in PBS buffer, followed by addition of 100 μL Chonblock Blocking/Sample Dilution (CBSD) ELISA buffer (Chondrex Inc, Redmond, USA). This was incubated at room temperature (RT) for 2 hours. Serum was tested at a dilution of 1:100 in CBSD ELISA buffer, then added to the wells for 2 hours at 37°C. After washing with PBS containing 0.1% Tween 20, horseradish peroxidase (HRP)-conjugated goat anti-human IgG (1:5,000) (GE Healthcare, Chicago, USA) was added for 1 hour at 37°C, followed by washing five times with PBS containing 0.1% Tween 20. HRP substrate (Ncm TMB One, New Cell & Molecular Biotech Co. Ltd, China) of 100 μL was added for 15 minutes, and the reaction was stopped by 50 μL of 2 M H_2_SO4. The OD was analysed in a Sunrise absorbance microplate reader (Tecan, Männedorf, Switzerland) at 450 nm wavelength. The background OD in PBS-coated control wells with the participant’s serum was subtracted from each OD reading. Values at or above an OD450 of 0.5 were considered positive and values below were imputed as 0.25 and considered negative.

*Surrogate virus neutralization test (sVNT)*

sVNT was conducted according to the manufacturer’s instructions (GenScript Inc, Piscataway, USA) and as described in our previous publication.^1,2^ The sVNT was performed using 10 μL of each serum, positive and negative controls, which were diluted at 1:10 and mixed with an equal volume HRP conjugated to the WT or Omicron BA.1 SARS-CoV-2 S-RBD (6 ng). The mixture was incubated for 30 minutes at 37°C, then 100 μL of each sample was added to microtitre plate wells coated with angiotensin-converting enzyme-2 (ACE-2) receptor. This plate was sealed for 15 minutes at 37°C and then washed with wash-solution, tapped dry, and 100 μL of 3,3',5,5'-tetramethylbenzidine (TMB) was added and incubated in the dark at RT for 15 minutes. This reaction was terminated using 50 μL of Stop Solution and the absorbance was read at 450 nm in a microplate reader. After confirming the positive and negative controls provided the recommended OD450 values, the % inhibition of each serum was calculated as (1 - sample OD value/negative control OD value) x100%. Inhibition (%) of at least 30%, the limit of quantification (LOQ), was regarded as positive, and values below 30% were imputed as 15%.

*T cell responses*

Peripheral blood mononuclear cells (PBMCs) were isolated from whole blood by density gradient separation then frozen in liquid nitrogen until use. Thawed PBMCs were rested for 2 hours in 10% human AB serum supplemented RPMI medium.

Next, the cells were stimulated with sterile ddH2O (as the peptide we ordered were dissolved with sterile ddH2O) or 1 µg/mL overlapping peptide pools representing the WT SARS-CoV-2 S, N and M proteins (Miltenyi Biotec, Bergisch Gladbach, Germany), or B.1.1.529/BA.1 S mutation pool and WT reference pool (Miltenyi Biotec, Bergisch Gladbach, Germany), omicron BA.1 N mutation pool, WT N reference pool, BA.1 M mutation pool and WT M reference pool (synthesized by ChinaPeptides Co., Ltd) for 16 hours in the presence of 1 µg/mL anti-CD28 and anti-CD49d costimulatory antibodies (clones CD28.2 and 9F10, Biolegend, San Diego, USA). After 2 hours of stimulation, 10 µg/mL brefeldin A (Sigma, Kawasaki, Japan) was added.^3^

| **Peptide sequences of N and M BA.1 mutation and WT reference pools** | | |
| --- | --- | --- |
|  | **BA.1 mutation** | **WT reference** |
| **N** | | |
| P13L | GPQNQRNAPRITFGG | GPQNQRNALRITFGG |
| 31_33delERS | GSNQNGERSGARSKQ | GSNQNGGARSKQ |
| 203_204delRGinsKR | STPGSSRGTSPARMA | STPGSSKRTSPARMA |
| **M** | | |
| D3G | MADSNGTITVEELKK | MAGSNGTITVEELKK |
| Q19E | LLEQWNLVIGFLFLT | LLEEWNLVIGFLFLT |
| A63T | WLLWPVTLACFVLAA | WLLWPVTLTCFVLAA |

The cells were then washed and subjected to immunostaining using a fixable viability dye (eBioscience, Santa Clara, USA, 1:60) and antibodies against CD3 (HIT3a, 1:60), CD4 (OKT4, 1:60), CD8 (HIT8a, 1:60), IFN-γ (B27, 1:15) and IL-2 (MQ1-17H12, 1:15) antibodies (Biolegend, San Diego, USA). Data acquisition was carried out using flow cytometry (LSR II; BD Biosciences, Franklin Lakes, USA) and analyzed by Flowjo v10 software (BD, Ashland, USA).

The antigen-specific IFN-γ^+^ or IL-2^+^ T cells were calculated by subtracting the background (sterile ddH2O) data, and presented as the percentage of CD4^+^ or CD8^+^ T cells.^4^ T cell response against a single peptide pool was considered positive when the frequency of cytokine-expressing cells was higher than 0.005% and the stimulation index was higher than 2; negative values were imputed as 0.0025%. Total T cell responses against S, N and M peptide pools were also added together, with a cut-off of 0.01%.

**SUPPLEMENTAL INFORMATION: SAFETY AND REACTOGENICITY**

**Adverse event: grade 2 tinnitus**

The patient (age range 11-18 years, female) who developed tinnitus post vaccination, had a history of a congenital anomaly of kidney and urinary tract (CAKUT) and underwent deceased donor kidney transplant 5 years ago. She was maintained on prednisolone, azathioprine, and tacrolimus. She developed tinnitus 8 days after dose 2 of 0.3ml BNT162b2 with left ear pain, left mid-facial numbness, back head pain, arthralgia, and tachycardia. The event was deemed to be possibly relevant to study vaccination and receipt of additional doses of BNT162b2 was deferred.

**Adverse event: relapse**

One patient (age range 11-18 years, male) with frequently relapsing steroid-resistant nephrotic syndrome was hospitalized for nephrotic syndrome which relapsed 28 days after the dose 3 of 0.3ml BNT162b2. He complained of increased puffiness with progressively increasing proteinuria since the day of dose 3. He was discharged after 28 days of hospitalization. Another patient with IgA nephropathy developed gross hematuria 1 day following dose 3 BNT162b2 and did not require hospitalization.

**Severe adverse events**

One patient (age range 11-18 years, male) had a history of lupus nephritis, currently on mycophenolate mofetil, prednisolone and hydroxychloroquine, was hospitalized for suspected myopericarditis 119 days after dose 3 of 0.3 ml BNT162b2. He was managed conservatively and was discharged 6 days with complete recovery, and his lupus disease activity remained well-controlled. The event was deemed to be not related to study vaccination due to lack of temporal association. The other two patients who experienced an SAE were a boy (age range 11-18 years) with autosomal recessive polycystic kidney disease on hemodialysis who was admitted for prolonged post-vaccine fever 6 days after dose 2 of 0.3ml BNT162b2, and a girl (age range 5-11 years) with ANCA-associated vasculitis on hemodialysis admitted for catheter-associated infection 31 days after dose 2 of 0.3ml BNT162b2 respectively.
